# Seroprevalence and Dynamics of anti-SARS-CoV-2 antibody among healthcare workers following ChAdOx1 nCoV-19 vaccination

**DOI:** 10.1101/2021.07.20.21260278

**Authors:** Soma Sarkar, Shantanab Das, Kabita Choudhury, Saibal Mukherjee, Raghunath Chatterjee

**Affiliations:** Department of Microbiology, NRS Medical College & Hospital, 138, A J C Bose Road, West Bengal, Kolkata, India 700014; NRS Medical College & Hospital, 138, A J C Bose Road, West Bengal, Kolkata, India 700014; Human Genetics Unit, Indian Statistical Institute, 203 B. T. Road, Kolkata, West Bengal, India 700108

**Keywords:** Immunoglobulin G, Seroprevalence, SARS-CoV-2 vaccine, COVISHILED, Neutralizing antibody, Anti-SARS-CoV-2 IgG Antibody stability

## Abstract

**Background:** The serological evaluations of IgG, IgM, and IgA to the SARS-CoV-2 proteins are widely used for the epidemiological assessment of COVID-19. The Health Care Workers (HCWs) are presumably exposed to a higher risk of acquiring the disease owing to their regular contact with the patients.

**Methods:** COVID-19 prevalence was investigated by classifying 313 HCWs into four groups based on their degree of exposure and estimating the IgG and total antibody. The serological assessment of the anti-SARS-CoV-2 antibody was conducted 21 days post-vaccination of first or both doses of the ChAdOx1 nCoV-19 vaccine among 174 HCWs. The vaccinated HCWs were followed up for 3 months for SARS-CoV-2 infection.

**Findings:** The levels of anti-SARS-CoV-2 IgG were comparable among different groups, but the seroprevalence gradually decreased from the most exposed to the less exposed group. The neutralizing antibody was positively correlated with IgG as well as total antibody. IgG was marginally decreased after 2 months followed by a significant drop after 4-6 months post-infection. However, 80% of the HCWs developed a detectable amount of IgG after the first dose of vaccination, the median titer of which was comparable to the seropositive HCWs after natural infection. Almost 100% of the HCWs developed antibodies after the second dose of vaccine with boosting effect among the seropositive HCWs. Although ∼11.5% of the vaccinated HCWs were infected with the SARS-CoV-2, ∼94% of them showed mild symptoms and recovered in home isolation without any O_2_ support.

**Interpretations:** The varying level of seroprevalence among the four groups suggested stratified spread of the disease. One dose of SARS-CoV-2 vaccination was found to be effective in terms of the antibody titer, while the second dose was required to cover the larger population. The effectiveness of the ChAdOx1 nCoV-19 vaccine was noticeable due to the low rate of post-vaccination infection with moderate or severe symptoms.

## Introduction

Coronavirus disease 2019 (COVID-19) started as a regional epidemic in December 2019 in Wuhan, China, but its rapid expansion made it a global pandemic affecting almost all the countries and significant mortality ^1,2^. While every affected country has taken containment and mitigation measures, the spread of COVID-19 is still prominent ^3^. The spectrum of clinical syndromes caused by SARS-CoV-2 ranges from asymptomatic cases to mild flu-like symptoms to severe pneumonia, acute respiratory distress syndrome (ARDS) and death. Many experts believe that unnoticed, asymptomatic cases of coronavirus infection could be the hidden source of contagion ^4-7^. The Health care workers (HCW) are the frontline workforce for clinical care and are presumably exposed to a higher risk of acquiring the disease than the general population. If infected, they not only pose a risk to the vulnerable patients and the fellow HCW ^8-10^ but associated morbidity and mental stress also cause disruption of patient care ^11^.

After primary infection, IgG antibody production can be maintained for a long time in any viral infection ^12^. The cell entry of SARS-CoV-2 is facilitated by interaction of the spike (S) glycoprotein through its receptor-binding domain (RBD) to the human ACE2 (hACE2) receptor and IgG antibody develops between 6-15 days following the disease-onset ^13^. The seroprevalence studies give us an indirect estimate of the proportion of HCWs who have experienced recent or past infection with COVID-19. Monitoring the prevalence of infection among HCW (regardless of history of symptoms) is useful for assessing the extent of exposure among the hospital personnel and identifying the high-risk groups. Considering the urgent need of vaccine across the globe, 120 vaccine candidates were reported within the first 5 months of 2020^14^. Many countries including India have initiated the vaccination drive^15^. Apart from clinical trial data more insights on antibody production are also needed for evaluation of the results of vaccination considering development of protective as well as therapeutic antibodies. Amidst the slow vaccination process in India, the number of cases significantly increased during the second wave ^16^.

Although there is evidence on the immunological responses against SARS-CoV-2, but the time to seroconversion and the antibody levels elicited in relation to the patient profile are not fully characterized yet. Importantly, the correlation between seropositivity or antibody levels and protection against re-infection, as well as the duration of protective immunity, remains to be elucidated.

The present study highlights several key aspects of antibody generation among HCWs in a tertiary care hospital of Kolkata. We determined the overall infection prevalence (past and current) to SARS-CoV-2, the prevalence of asymptomatic infections, SARS-CoV-2 antibody kinetics and longevity as well as the development of protective antibody in HCWs following infection. Additionally, we also investigated the level of neutralizing antibody and its relationship with the IgG and total antibody among seroreactive individuals. The levels of antibody production after first and second doses of vaccination were also investigated among the HCWs. The rate of infection among the vaccinated HCWs within 3 months post-vaccination and the degree of severity and hospitalization in case of infection were also evaluated.

## Materials and Methods

### Study setting

This cross-sectional study among the health care workers (HCWs) was executed at a tertiary care hospital, Kolkata after obtaining approval from the Institutional Ethics Committee, Nilratan Sircar Medical College & Hospital. Among the 3,997 HCWs in the tertiary care center, 313 HCWs (11 Administrative staff, facility manager and clerical staff, 66 Junior doctors, 38 Medical officer, 81 Nursing staff, 24 Paramedical staff and research scientist, 93 Support staff, GDA, security, kitchen staff) were randomly selected and included in the study with their informed consent for serosurvey during November 2020 to January 2021. The follow up and antibody response studies following vaccination were conducted during January 2021 to June 2021.

The study population were chosen randomly irrespective of their working department and divided into 4 groups (A, B, C and D) based on their working area, direct care to COVID-19 patient, face-to-face contact (within 1 meter) and duration of exposure to the COVID patients, performed any aerosol-generating procedures or direct contact with the environment where the confirmed COVID-19 patient was cared like bed, linen, medical equipment, bathroom etc.^17^. Individuals within Group A were the most exposed and Group D were the least. HCWs with 5 hours of cumulative exposure per day to the covid patient directly or indirectly were included in Group A. Those HCWs with 4hrs, 3hrs and 1hr cumulative exposure per day to the COVID-19 patient were included in Group B, Group C and Group D, respectively. Participants with symptoms suggestive of recent infection or positive RT-PCR test result within last 14 days were excluded from the study. A structured questionnaire comprising demographics, prior symptoms with their duration, prior COVID-19 test results, working location (in terms of working in COVID or Non-COVID unit), co-morbidities were collected prior to the blood collection for SARS-CoV-2 antibody testing.

### Serological analysis of IgG and Total Antibody

Serum samples were prepared from the clotted blood following centrifugation for 10 minutes at 3000g at room temperature. Serological analysis of IgG and total antibody (IgG, IgM and IgA) were performed using enhanced chemiluminescence technology by Vitros ECiQ (Ortho Clinical Diagnostics, New Jersey, US). In this immuno-metric technique, antibodies to SARS-CoV-2, if present in the sample bind to the SARS-CoV-2 spike protein coated on the wells. When horseradish peroxidase (HRP) labelled murine monoclonal anti-Human IgG antibodies were added as conjugate, the conjugate interacted specifically to the antibody portion of antigen-antibody complex which is measured by a luminescent reaction. After the addition of luminogenic substrate, HRP bound conjugate catalyzes the oxidation of the luminal derivative, producing light. The electron transfer agent (substituted Acetanilide) increases the level of light and prolongs its emission. The amount of HRP conjugate bound is directly proportional to the amount of SARS-CoV-2 antibody present in the sample ^18^.

### Neutralizing antibody sandwich ELISA

To find out whether the seropositive patients were also developing the neutralizing antibody, a neutralizing antibody sandwich ELISA (GenScript, USA) following manufacturer’s protocol was also performed. SARS-CoV-2 spike protein contains a receptor binding domain (HRP-RBD), which recognizes the human ACE2 receptor (hACE2) protein on the cell surface and leads to endocytosis into the host cell. This neutralization assay mimics the virus neutralization process through virus-host interaction. The diluted samples, if contains neutralising antibody, bound with HRP-RBD and the protein-protein interaction between HRP-RBD and hACE2 is blocked by neutralising antibody (blocking ELISA) ^19^.

### Dynamics of IgG antibody over time

Among 313 HCWs, 104 were RT-PCR confirmed COVID-19 patients. To evaluate the dynamics of the antibody titer, all RT-PCR confirmed COVID-19 HCWs were followed up at two months interval for six months of their first antibody measurement.

### Seroreactivity after vaccination

247 HCWs, who received ChAdOx1, nCoV-19 corona virus vaccine (COVISHIELD), a replication deficient simian adenoviral vector that expresses full length Spike (S) protein of SARS-CoV-2 ^20^, were included in the study. Blood samples were collected from 130 HCWs twenty-one days after first dose and from 76 HCWs twenty-one days after second dose (32 HCWs were common in both doses) of ChAdOx1, nCoV-19 vaccine for anti-SARS-CoV-2 antibody testing.

### Statistical Analyses

Descriptive as well as inferential analyses were performed using R Software ^21^. A significance level of p ≤ 0.01 was considered unless it was specifically mentioned.

## Results

The mean age of the 313 HCWs was 37.83 (SD=12.31) with 54% male and 46% female. Around 51% of them were involved in direct care of the COVID-19 patients with different level of exposures. Based on their occupational risks and degree of exposures, HCWs were classified into four groups. The mean ages of all four groups were also approximately similar. Besides their co-morbidities, COVID-19 like symptoms, working in COVID unit and use of prophylactic drug was also recorded for each subject (Table 1).

**Table 1:** Demographic details of study participants.

|  |  | ALL (n=313) | Group A (n=104) | Group B (n=81) | Group C (n=66) | Group D (n=62) |
| --- | --- | --- | --- | --- | --- | --- |
| **Sex** | **Male** | 168 | 83 | 1 | 35 | 49 |
|  | **Female** | 145 | 21 | 80 | 31 | 13 |
| **Age** | **Median** | 36 | 38 | 40 | 26 | 41 |
|  | **Mean (SD)** | 37.83 (12.31) | 39.71 (12.39) | 40.6 (12.74) | 26.89 (3.68) | 42.72 (11) |
| **Working in COVID Unit** |  | 161 | 35 | 45 | 43 | 38 |
| **No underlying illness** |  | 244 | 81 | 56 | 61 | 46 |
| **HCQ Prophylaxis taken** |  | 86 | 12 | 21 | 19 | 34 |
| **Symptomatic** |  | 119 | 48 | 27 | 25 | 19 |

### Serological analysis of IgG and Total Antibody

Around 34% (106) of the HCWs were seropositive for IgG while 40% (126) of them were seropositive for total antibody (IgG, IgM and IgA). The median IgG titer (signal to cut-off ratio, henceforth referred as S/Co ratio) was 6.30 with median absolute deviation (MAD) was 3.27 for IgG reactive individuals. Whereas the median total antibody titer was 141 S/Co ratio (MAD=101.60) among seroreactive HCWs. Seroprevalence was not significantly different with respect to age or sex of the HCWs (p-value=0.4798, OR=1.18, 95% CI=0.74-1.88). Although the median IgG titer were comparable for four groups (median S/Co ratio for Group A = 5.48 and MAD = 3.65, Group B = 6.08 and MAD =3.49, Group C = 6.22 and MAD=2.68 and Group D = 7.89 and MAD =2.86), but the seroprevalence was higher in Group A (53.8%) and gradually decreased for Group B (40.7%), Group C (31.8%), and Group D (25.8%) for total anti-SARS-CoV-2 antibody (Figure 1a). Whereas for IgG, Group A (38.4%) and Group B (39.5%) showed similar seroprevalence and significantly lower prevalence in Group C (30.3%), and Group D (22.5%) (Figure 1A). We further investigated the total antibody titers among IgG reactive and non-reactive individuals. The level of total antibody among IgG reactive HCWs were significantly higher compared to the level of IgG, suggesting presence of IgM and IgA (Figure 1B). Total antibody among IgG non-reactive HCWs varied from 0 to 140 S/Co ratio (Figure 1C). Around 35% HCWs, who worked in the COVID-19 unit were seropositive. A similar proportion of seropositive HCWs were also observed in non-COVID unit HCWs (p-value=0.9154, OR=1.08, 95% CI=0.64-1.63). Following ICMR guidelines^22^, HCWs were advised to consume hydroxychloroquine (HCQ) as a prophylactic medication. In our entire study population only 27% have consumed HCQ as prophylaxis. The seropositivity among HCQ consumer and non-consumer was not significantly different (p-value=0.3285, OR=0.776, 95% CI=0.44-1.30) (Table 2). As consumption of HCQ did not significantly affect the entire population, we didn’t pursue it for further analysis on group wise effect of HCQ.

**Table 2:** Association of COVID-19 symptoms with SARS-CoV-2 infection.

| **Total Sample** |  |  |
| --- | --- | --- |
| **Factors** | **Odds Ratio (95% CI)** | **P value** |
| **Sex** | 1.18 (0.74-1.88) | 0.4798 |
| **COVID-19 Suggestive Symptoms** | **3.49 (2.14-5.70)** | **3.05E-07** |
| **Fever and Headache** | **3.39 (2.05-5.62)** | **1.18E-06** |
| **GI upset** | **2.88 (1.06-7.81)** | **0.03035** |
| **Cough and Sore throat** | **2.24 (1.26-3.97)** | **0.005051** |
| **HCQ Prophylaxis** | 0.77 (0.44-1.30) | 0.3285 |

**Figure 1.**
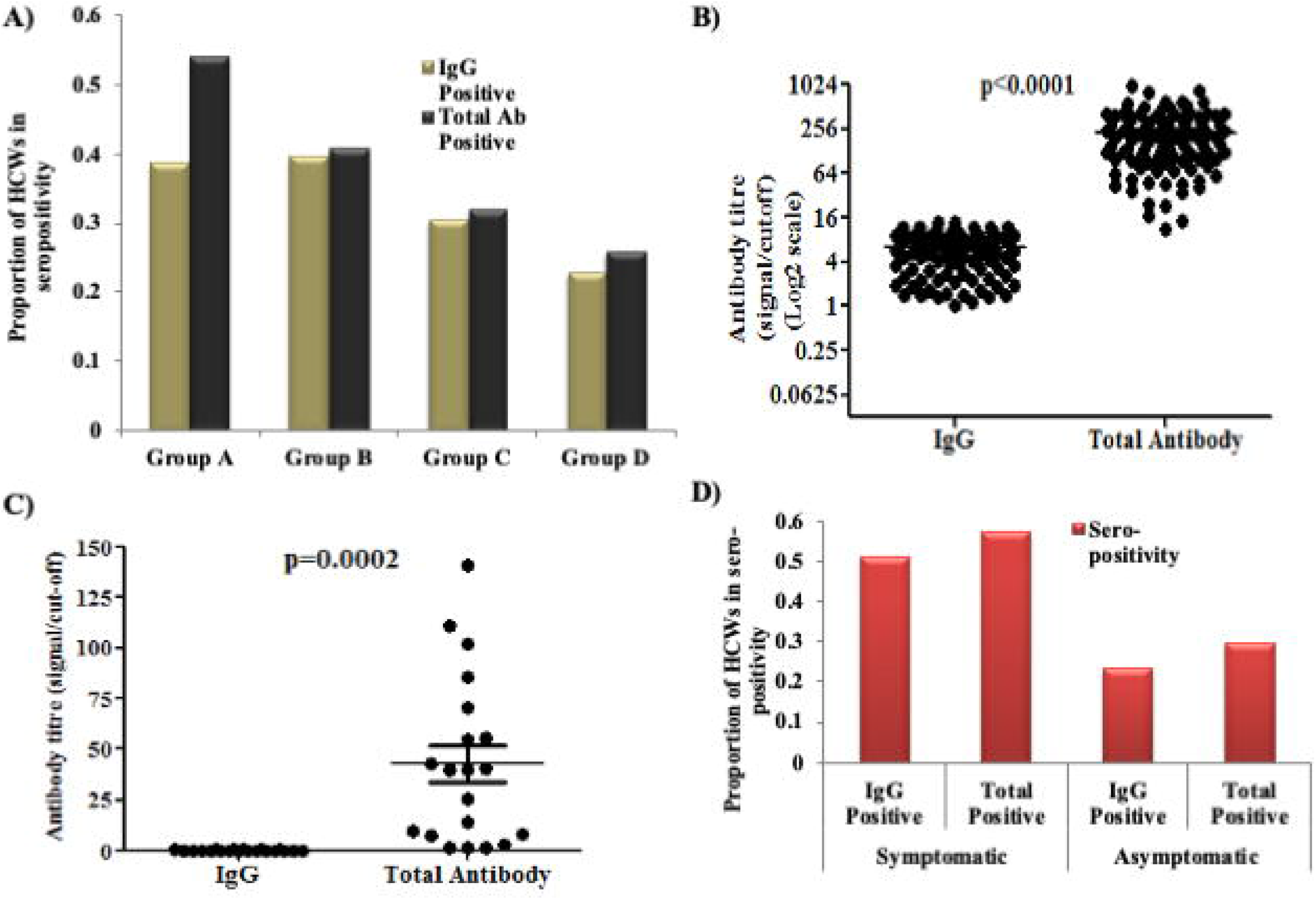
Seroprevalence among health care workers. **(A)** Seroprevalence using anti-SARS-CoV-2 IgG and total antibody in four groups of HCWs. **(B)** Signal/cut off ratio of IgG and total antibody of IgG positive HCWs. **(C)** Signal/cut off ratio of IgG and total antibody of IgG negative HCWs. **(D)** Seroprevalence among COVID-19 suggestive symptomatic and asymptomatic individuals. Around 25% more symptomatic individuals developed antibody than asymptomatic individuals.

Next, we wanted to evaluate the IgG prevalence among symptomatic and asymptomatic HCWs. Around 38% of the studied population developed at least one of the COVID-19 symptoms in recent past. We identified 24% of the asymptomatic HCWs developed IgG, whereas 52% of the symptomatic individuals were found to be seropositive for SARS-CoV-2 antibody (OR=3.49, 95% CI=2.14-5.70, p-value=3.5×10^−7^) (Figure 1D). We further evaluated the association of seropositivity among different COVID-19 suggestive symptoms, and found a significant association with all symptoms, among which fever and headache showed higher significance compared to other symptoms (OR=3.397, 95% CI=2.05-5.62, p-value=1.179×10^−6^) (Table 2).

### Neutralizing antibody sandwich ELISA

To assess the extent of neutralizing activity of the seropositive HCWs, we randomly selected 46 individuals from the IgG seroreactive group and determined the level of neutralizing antibody. The level of neutralizing antibody was found to be positively correlated with the IgG S/Co ratio (R^2^=0.8363, p-value<0.001) (Figure 2A) suggesting that the IgG seropositive HCWs developed the protective antibody against SARS-CoV-2.

**Figure 2.**
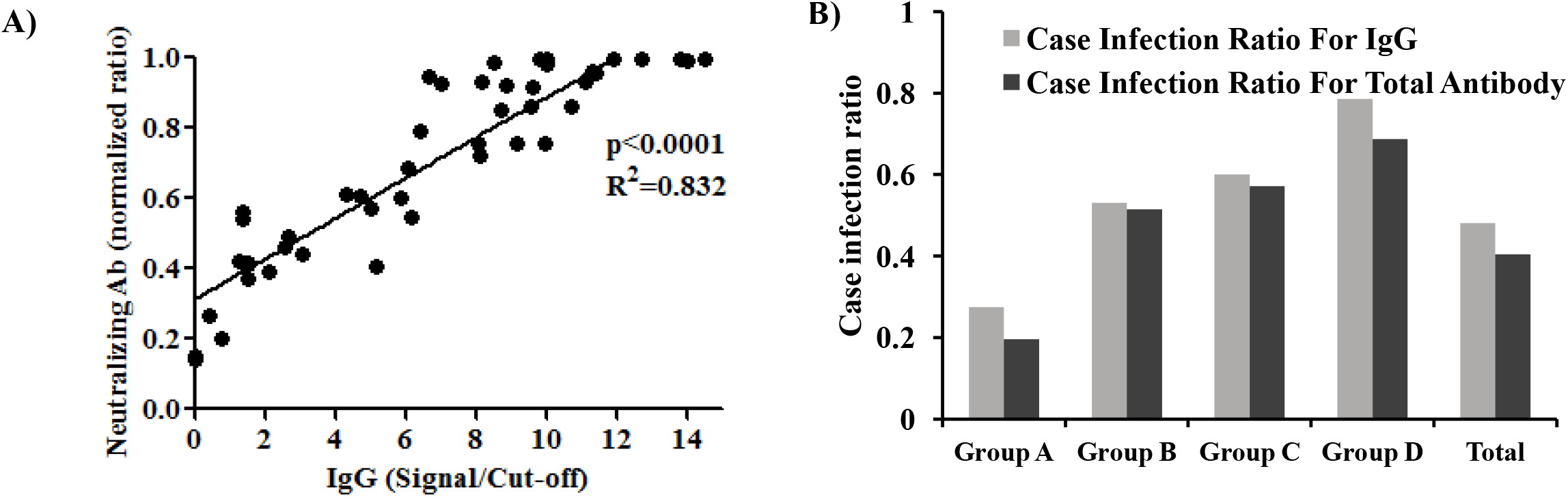
**(A)** Correlation between IgG and neutralizing antibody titer (S/Co ratio). The line graph shows significant positive correlation between IgG and neutralizing antibody titer. **(B)** Case infection ratio among four occupational risk groups and all (total) samples.

### Case infection ratio among HCWs

We further analysed the case detection using RT-PCR among the four groups of seropositive HCWs. Around 84% of RT-PCR positive HCWs showed detectable IgG, whereas 92% of the HCWs showed detectable level of total antibody. We determined the case-infection ratio for each group, where case refers to the RT-PCR tested COVID-19 positive cases and infection refers to the seroreactive individuals in each group. Among the total samples, we found a case-infection ratio of 0.48 for IgG, suggesting that around half of the HCWs did not opt for the RT-PCR test (Figure 2B). In group wise analysis, a gradual increase of case-infection ratio from Group A to Group D was observed for both IgG and total antibody (Gr A= 0.27, B=0.53, C=0.60 and D=0.78).

### Dynamics of anti-SARS-CoV-2 IgG antibody over time

To evaluate the antibody titer over time, we classified the RT-PCR confirmed COVID-19 HCWs with respect to time duration of COVID-19 RT-PCR testing to the first IgG measurement. The IgG and total antibody titers were found to be decreasing with times. The median value of IgG S/Co ratios were found to be 7.75 (MAD=2.77), 5.97 (MAD=3.21) and 1.70 (MAD=1.685) at 0-2 months, 2-4 months, and 4-6 months, respectively. Both IgG and total antibody titers were found to be significantly decreasing at 4-6 months post-infection (p=0.04) (Figure 3A, B). To get more insight about this trend, we classified HCWs into three groups having IgG titers of 1-3, 3-6 and >6 S/Co ratio and estimated the proportion of individuals within each group. Around 69% of HCWs showed IgG > 6 S/Co ratio between 0-2 months and started decreasing at 2-4 months followed by a significant drop at 4-6 months post-infection (Figure 3C). To substantiate this observation, we performed a follow up study with randomly selected 42 IgG seropositive individuals at 2 months interval. The IgG titer at 0-2 months showed a significant decrease at 2-4 months (p=0.002) indicating the significant decreasing trend of IgG over time (Figure 3D).

**Figure 3.**
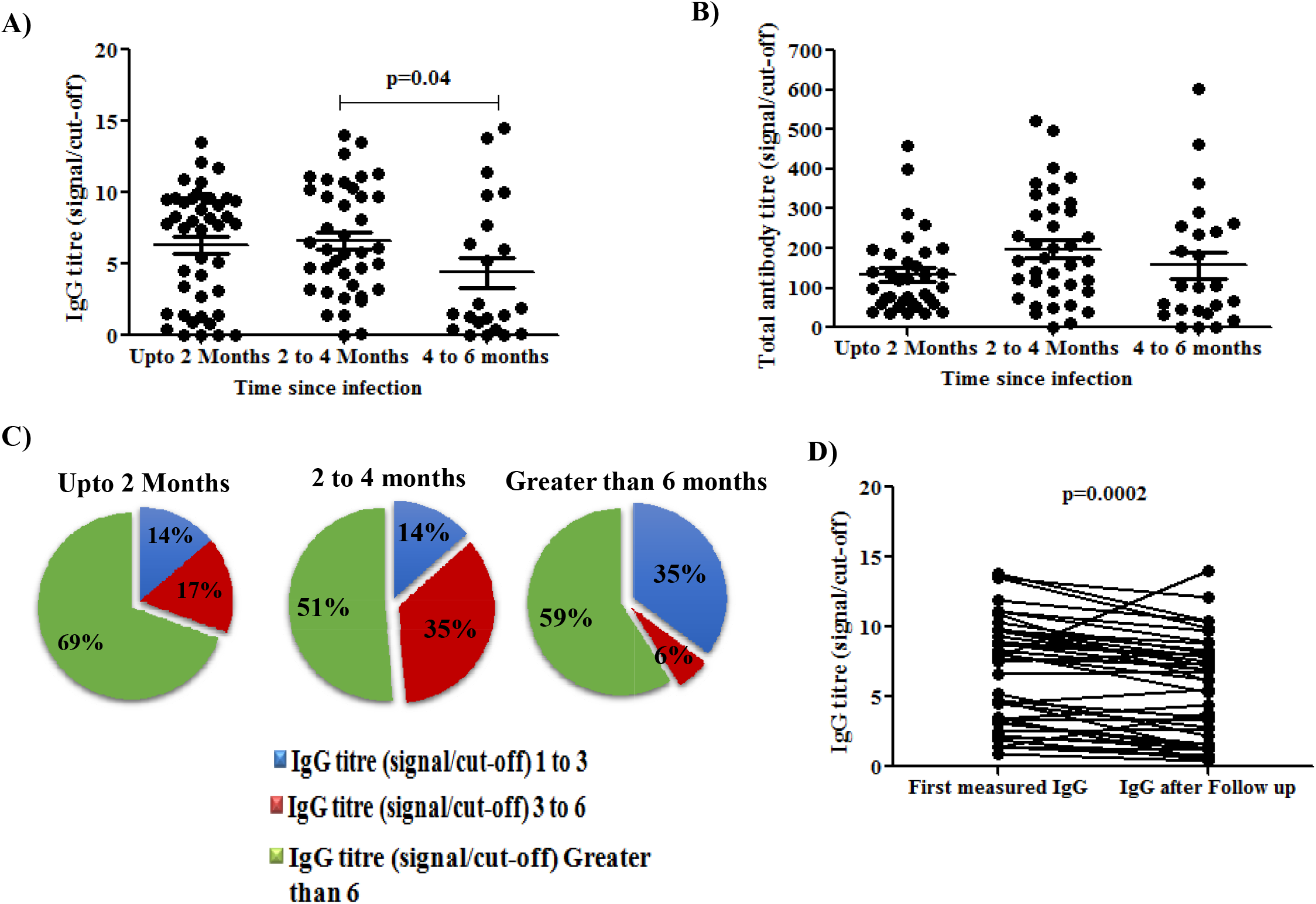
IgG dynamics among seropositive HCWs with time. **(A)** IgG titer measured at 0-2 months, 2-4 months and 4-6 months post infection among RT-PCR confirmed COVID-19 cases. **(B)** Total antibody titer measured at 0-2 months, 2-4 months and 4-6 months post infection among RT-PCR confirmed COVID-19 cases. **(C)** Pie chart representing proportion of HCWs with IgG S/Co ratio at 0-2 months, 2-4 months and 4-6 months post infection among RT-PCR confirmed COVID-19 cases. **(D)** IgG titer (S/Co ratio) and the same after following up at two months interval.

### Seroreactivity after vaccination

Next, we evaluated the effect of vaccination on development of anti-SARS-CoV-2 IgG. IgG titers of 130 HCWs after first dose and 76 HCWs after second dose of ChAdOx1 nCoV-19 vaccine were determined. Around 80% of the HCWs, who has taken first dose have developed detectable IgG, but surprisingly remaining 20% of them did not develop detectable amount of antibody against SARS-CoV-2 after 21 days from the first dose of vaccine (Figure 4A). To better understand the antibody response among these non-reactive individuals, we categorized the first dose post-vaccination HCWs according to previous IgG status. It was observed that the IgG titer was significantly enhanced to 14.7 S/Co ratio (MAD=0.9) after first dose of vaccination compared to previously seropositive (7.18 S/Co ratio (MAD=3.12)) HCWs (p<0.0001) (Figure 4B). In case of previous seronegative HCWs, the median IgG was found to be 7.26 S/Co ratio (MAD=2.74) after first dose of vaccination (Figure 4C). Notably, the RT-PCR confirmed COVID-19 HCWs showed a median IgG titer of 8.25 S/Co ratio (MAD=1.38) within 1-2 months post infection, indicating comparable effect of the first dose of vaccination with that of the natural infection by SARS-CoV-2 (Figure 4D). This observation clearly indicated that the first dose of vaccine among previously seropositive individuals acted as booster to enhance the level of antibody.

**Figure 4.**
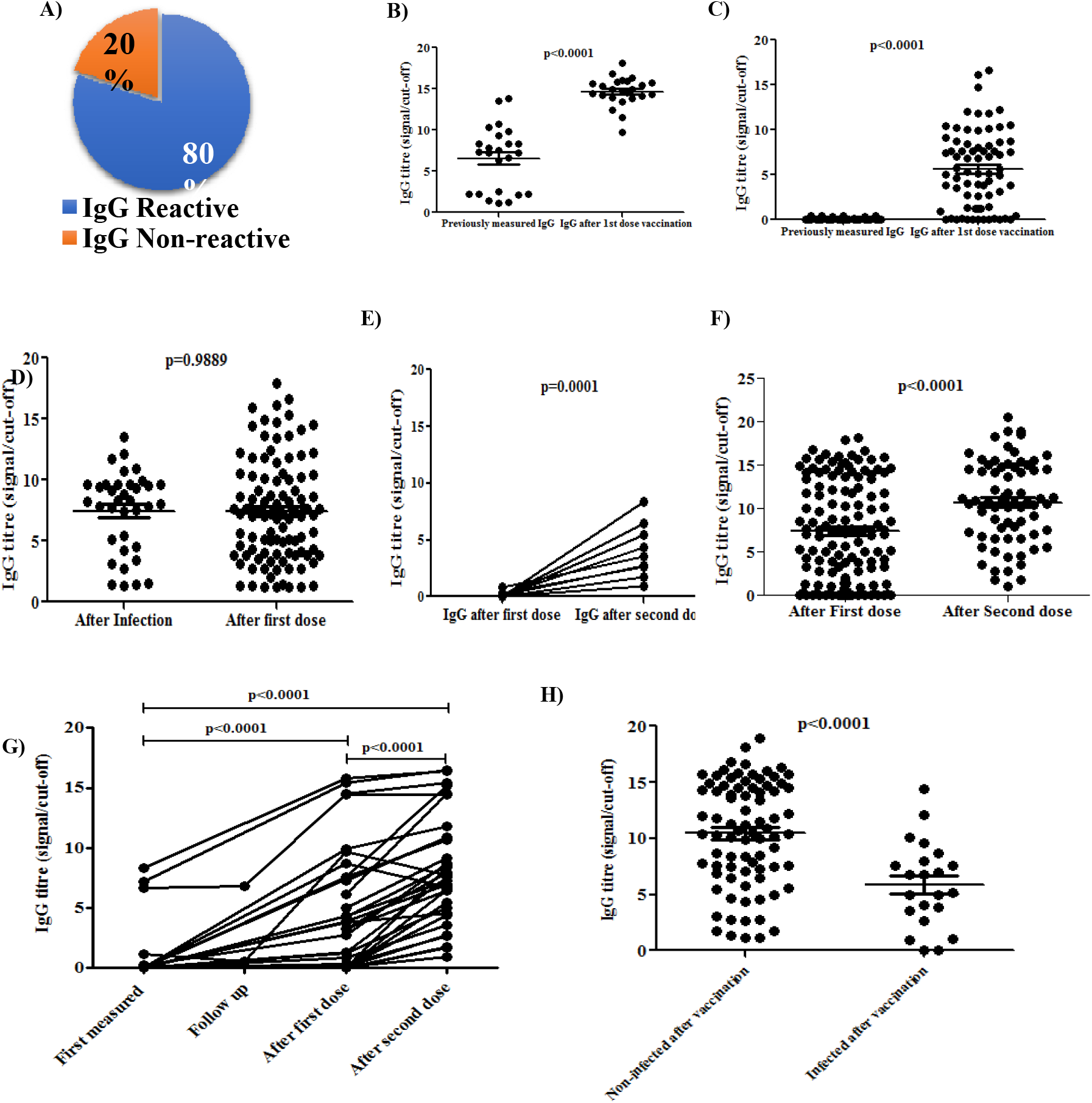
**(A)** Pie chart representing proportion of IgG reactive and non-reactive HCWs after first dose of vaccination. **(B-C)** IgG titer (S/Co ratio) among previously IgG B) seropositive HCWs after first dose of vaccination and C) seronegative HCWs after first dose of vaccination. **(D)** IgG titer (S/Co ratio) among naturally infected IgG seropositive HCWs and seronegative HCWs after first dose of vaccination. **(E-F)** IgG titer at 21 days post first and second dose of vaccines among E) seronegative and F) seropositive HCWs after first dose vaccination. **(G)** The longitudinal data of IgG titers (S/Co ratio) from first measured to second dose of vaccination. **(H)** IgG titer (S/Co ratio) among the vaccinated HCWs with or without SARS-CoV-2 infection within 3 months post vaccination.

Most of the vaccination strategy used a two-dose programme for the complete vaccination. To evaluate the antibody dynamics following two doses of vaccination, we determined the IgG of 130 HCWs after first dose and 76 HCWs after second dose of vaccine. Twenty-one days post first dose of vaccination, we found ∼20% HCWs remained seronegative while ∼100% of them become seropositive after 21 days post second dose of vaccination, suggesting importance of second dose of vaccine (Figure 4E). We further classified the HCWs as first dose non-reactive and reactive groups and determined antibody titer following 2 doses of vaccination. Interestingly, the non-reactive individuals after first dose of vaccination became seropositive to anti-SARS-CoV-2 IgG after the second dose (p=0.0001) (Figure 4E), whereas the IgG titer after second dose (median=10.7, MAD=3.8) significantly enhanced compared to first dose (median=7.26, MAD=5.9) (p<0.0001) (Figure 4F). Longitudinal data of anti-SARS-CoV-2 antibody replicated the general observation of significant enhancements after both first and second doses of ChAdOx1 nCoV-19 vaccine compared to the antibody titer before vaccination (Figure 4G).

In our cohort of 313 HCWs, 247 HCWs received both doses and 19 received only first dose of the ChAdOx1 nCoV-19 vaccine until June 2021. Among 266 ChAdOx1 nCoV-19 vaccinated HCWs, 31 (11.5%) were infected with SARS-CoV-2 within three months post vaccination. Interestingly, 94% of the infected HCWs showed either mild or no symptoms and recovered in home isolation without requirement of any O_2_ support. Only 1 HCW needed hospitalization with severe symptoms and O_2_ requirement. To evaluate whether the level of antibody could serve as a possible indicator of post vaccination infection, we compared the median antibody titer among the post-vaccination uninfected and infected HCWs. The median antibody S/Co ratio for the infected HCWs (median=5.94, MAD=2.225) was significantly less compared to the uninfected HCWs (median=10.7, MAD=3.8) (p<0.001) (Figure 4H), suggesting a possibility of SARS-CoV-2 infection among the low antibody titer seropositive individuals. Note that other factors such as degree of precautionary measures, occupational risk factors among these groups might also be a perilous factor for such infection.

## Discussion

To understand the transmission dynamics of SARS-CoV-2 virus, seroprevalence is important to guide interventions for control of COVID-19 pandemic. Beside the RT-PCR based detection, the seroprevalence acts as an important measure of the potential past infection for mild and asymptomatic patients. In a tertiary care hospital set up, seroprevalence guides the COVID-19 infection dynamics among the different working groups ^23^. As HCWs are the most exposed to SARS-CoV-2 infection, determination of seroprevalence among different working groups is an effective indicator to monitor and control the spread of SARS-CoV-2 infection. Seroprevalence rate was found to be different among HCWs in several countries due to various factors like selection bias in recruiting study participants, study period, awareness and effective implementation of infection control practices, adequate use of PPE, maintenance of hand hygiene and physical distance, early identification and timely isolation and quarantine as well as seroprevalence rate in the community ^24-28^.

In this study, the seropositivity among HCWs in a tertiary care hospital was 34% for IgG during November 2020 to January 2021, which was similar to a study conducted during April 2020 among the HCWs at a tertiary hospital in New York city, USA (36%) ^29^. It was observed that the seroprevalence among HCWs in this study was higher to the reported seroprevalence of 26% among the HCWs in Kolkata during September 2020 ^28^. The third pan India serosurvey, conducted during December 2020-January 2021 also reported 25.6% seroprevalence among the HCWs in India ^30^, suggesting variable seroprevalence among the HCWs across different parts of the country. In contrary to our observation, the pan India serosurvey did not identify any difference in seroprevalence between different HCW categories. This difference might be due to inadequate stratification of risk groups among the HCWs. However, variable prevalence among HCWs across different parts of the country might also impact the results of their study.

The overall seroprevalence data showed no significant association with age or sex, as also was observed in second and third pan India serosurvey conducted in August and December 2020 ^30,31^. As Group A and Group B were mostly exposed to COVID-19 patients, seropositivity of these two groups was similar, whereas Group C and Group D showed significantly low seropositivity compared to groups A and B. The decreasing trend of seroprevalence ranging from 54% to 23% from high risk to low-risk group for both IgG and total antibody vividly portrayed the association of seroprevalence with SARS-CoV-2 infection risk. The total antibody titer (S/Co ratio) was significantly higher than IgG because it includes IgA, IgM along with IgG. Among the IgG non-reactive individuals, 10% showed presence of detectable amount of total anti-SARS-CoV-2 antibody, suggesting presence of only IgA and IgM. The pan India serosurvey also reported variable seroprevalence in urban and rural areas with higher prevalence in urban slum areas than rural areas ^31^.

Working in COVID-19 or non-COVID-19 unit was not associated with increased antibody positivity in HCWs. The seroprevalence study from Spanish hospital also found no association of COVID-19 with working in COVID unit suggesting that awareness and strict adherence to infection control protocols were sufficient to prevent transmission to HCWs ^25^.

This study also evaluated the effect of HCQ as prophylaxis for COVID-19 in terms of seroprevalence among the HCQ consumers and non-consumers. In a hospital based seroprevalence study in Kolkata, there was a significant association of seroreactivity with adequate HCQ consumption as only 1.3% of HCQ consumers became reactive ^28^. But in this study, we did not find any effect of HCQ prophylaxis on seroprevalence of COVID-19.

We know that the cellular immune response plays an important role to clear the virus from host cell and humoral immune response is responsible for preventing future infection. Although the plaque reduction neutralization test (PRNT) is the gold standard for neutralization assays, it is cumbersome, time consuming and require Biosafety Level 3 facilities. Therefore, we performed ELISA based neutralizing assay to substantiate the preventive immunity of anti-spike IgG and found 100% of seropositive individuals developed detectable neutralizing antibody. The signal cut-off values of detected IgG were directly proportional to the level of neutralizing antibody.

We found a positive correlation of both IgG and total antibody reactivity with COVID-19 suggestive symptoms developed throughout the course of the disease as 52% of all symptomatic individuals were seroreactive. So, early identification of suggestive symptoms acts as a predictive indication of SARS-CoV-2 infection and based on that self-isolation can be recommended to prevent the further spread. Seroprevalence study at Sweden showed that almost all COVID-19 symptoms were highly associated the seroreactivity ^26^. The first seroprevalence study at Belgium also found significant association of COVID-19 symptoms with seroprevalence ^23^.

Detection of RNA of SARS-CoV-2 by RT-PCR or humoral responses to the virus (detectable IgG) is the evidence of SARS-CoV-2 infection. Here we determined the RT-PCR positive cases among IgG positive HCWs as case infection ratio for each group in our cohort. The gradual increasing trend of case infection ratio from group A to group D decisively showed that most of the infections were undetected in HCWs from high-risk groups whereas the low-risk group HCWs were aware of their infection and did RT-PCR test in time.

On the other hand, the durability of the antibody titer depends on its initial titer and the time of measurement following infection or vaccination. We found that IgG titer was significantly reduced after 4 to 6 months of infection and the proportion of individuals in each IgG titer level decreased over time. A brief follow-up after 2 months, with randomly selected 42 seropositive individuals also substantiated the decay kinetics of IgG. This data was concomitant with the study by Wajnberg et al., where it was shown that the titer could last up to 5 months ^32^. A CDC report on November 2020 showed similar observations where 94% of 156 seropositive HCWs experienced a rapid decline in antibody titer after 60 days of initial observation ^33^.

The remarkable volume of knowledge accumulated from the scientific quest during this pandemic helped in yielding actionable insights which lead to develop vaccines and therapeutic strategies against COVID-19. As of now, 102 vaccine candidates are at various stages of clinical trials, and 184 vaccines are in preclinical development spanning diverse vaccination platforms ^14^. Administration of adenovirus-based vaccines COVISHEILD was initiated in India for HCWs in early January, 2021 ^15^. We systematically analysed the development of anti-spike glycoprotein IgG antibody after first and second dose of COVISHIELD vaccination. Startlingly, 20% of them did not develop detectable anti-spike IgG after 21 days of the first vaccination. We also found that, for previously seropositive HCWs, the first dose acted as a booster dose and the detectable IgG titer was significantly elevated than previous antibody level. In contrary, seronegative individuals also developed detectable IgG after the first dose. Although, there was a reduction in IgG titer in pre-vaccinated follow up measurements, it showed a significant increase after first dose of vaccination. A comparable median IgG (8.26 S/Co ratio, MAD=1.38) were observed among the RT-PCR tested positive and the previously seronegative first dose vaccinated HCWs (7.35 S/Co ratio, MAD=3.06) suggested effectiveness of first dose vaccination for antibody production. After administration of the second dose, ∼100% of first dose seronegative individuals developed detectable IgG and the antibody titer was also significantly elevated. For the first dose seropositive individuals the IgG titer after the second dose was also significantly elevated. Exposure to the SARS-CoV-2 among the HCWs are generally higher compared to other individuals due to their involvement in patient care, especially in a COVID-19 tertiary care center. SARS-CoV-2 infections among the vaccinated HCWs was found to be ∼12%, while the disease severity among them was very low compared to the unvaccinated HCWs. Although, in our study we did not investigate the protection of antibody after vaccination on newly emerged SARS-CoV-2 variants, the recent report suggest that the vaccination with ChAdOx1 nCoV-19 is as effective against the B.1.1.7 variant of SARS-CoV-2 as other lineages and results in reduction in viral load as well as duration of virus shedding, which obviously decrease the transmission of disease ^34^. ChAdOx-1 nCov-19 vaccine showed 9-fold lower *in vitro* neutralization activity against B.1.1.7 (double mutant strain) versus a canonical non-B.1.1.7 strain ^34^. This observation along with the enhanced antibody generation supports the immense potential of ChAdOx-1 nCov-19 vaccine. To conclude, our study, which dealt with the anti-SARS-CoV-2 antibody dynamics starting from prevalence through follow-up up to the second dose of vaccination and its association on several significant factors, may help to build better preventive strategies in future. Similar comprehensive study over the general population following vaccination will be necessary to monitor the trend and optimal resource utilization for better management of the ongoing pandemic in a large country like India.

## Data Availability

Not applicable

## Ethical clearance

The study was approved by the Institutional Ethics Committee, Niratan Sircar Medical College & Hospital, 138 A. J. C. Bose Road, Kolkata 700104.

## Author contributions

SS, SM and RC conceptualize the study. SS, SD and KB performed the recruitment of participant, sample collection and experiments. SS, SD and RC performed the data analysis and wrote the manuscript.

